# Automatically Identifying Childhood Health Outcomes on Twitter for Digital Epidemiology in Pregnancy

**DOI:** 10.1101/2022.11.01.22281813

**Authors:** Ari Z. Klein, José Agustín Gutiérrez Gómez, Lisa D. Levine, Graciela Gonzalez-Hernandez

## Abstract

Data are limited regarding associations between pregnancy exposures and childhood outcomes. The objectives of this preliminary study were to (1) assess the availability of Twitter data during pregnancy for users who reported having a child with attention deficit/hyperactivity disorder (ADHD), autism spectrum disorders (ASD), delayed speech, or asthma, and (2) automate the detection of these outcomes. We annotated 9734 tweets that mentioned these outcomes, posted by users who had reported their pregnancy, and used them to train and evaluate the automatic classification of tweets that reported these outcomes in their children. A classifier based on a RoBERTa-Large pretrained model achieved the highest F_1_-score of 0.93 (precision = 0.92 and recall = 0.94). Manually and automatically, we identified 3806 total users who reported having a child with ADHD (678 users), ASD (1744 users), delayed speech (902 users), or asthma (1255 users), enabling the use of Twitter data for large-scale observational studies.

## INTRODUCTION

Many children are diagnosed with disorders that can impact their daily life and can last throughout their lifetime. For example, in the United States, 17% of children are diagnosed with a developmental disability [1], and 8% of children are diagnosed with asthma [2]. The causes of these disorders are likely multifactorial and largely unknown. Meanwhile, sources of data for assessing the association of these outcomes with pregnancy exposures remain limited, given that clinical trials typically exclude people who are pregnant [3], and pregnancy registries typically follow infants for up to one year after birth [4]. Despite attention deficit/hyperactivity disorder (ADHD) being one of the most common developmental disabilities, with a prevalence of 9% among children in the United States [1], and medication use occurring in approximately 90% of pregnancies [5], systematic and scoping reviews [6-9] have identified only a small number of cohort studies assessing associations between medication exposure during pregnancy and ADHD in the children.

Our recent work [10-12] demonstrated that Twitter data can be used to assess associations between maternal exposures (including in the periconception period) and miscarriage, preterm birth, low birthweight, birth defects, and neonatal intensive care unit (NICU) admission. While our previous work focused on prenatal, perinatal, and postnatal outcomes, our ability to continue collecting users’ tweets on an ongoing basis after birth [13] may present opportunities to detect outcomes in childhood. Twitter data has been used to identify self-reports of ADHD [14], autism spectrum disorders (ASD) [15], and asthma [16], but, to the best of our knowledge, it has not been used to identify reports of these disorders in users’ children. The objectives of this preliminary study were to (1) assess whether there are users who were active on Twitter during pregnancy and also posted tweets that reported having a child with ADHD, ASD, delayed speech, or asthma, and (2) develop and evaluate a natural language processing (NLP) method to automatically identify these outcomes in children for large-scale observational studies.

## METHODS

### Ethical considerations

The data used in this study were collected and analyzed in accordance with the Twitter Terms of Service. The Institutional Review Board of the University of Pennsylvania reviewed this study and deemed it exempt human subjects research under 45 CFR §46.101(b)(4) for publicly available data sources.

### Data collection and annotation

In recent work [17], we automatically identified more than 100,000 Twitter users who posted tweets self-reporting an ongoing pregnancy. For this study, we searched these users’ timelines—all of their tweets posted over time—for mentions of ADHD, ASD, delayed speech, and asthma, and their lexical variants [18], including misspellings. Our search also required the tweets to contain references to a child (e.g., *daughter, son, baby, child, kid, toddler, # year old*) and the user (e.g., *I, my, mother, parent*), but excluded tweets that matched specific patterns indicating the user’s own disorder (e.g., *I have autism, my asthma*). Excluding retweets, we identified a total of 36,094 matching tweets, posted by 11,712 users. Among these 36,094 matching tweets, 5823 tweets (2746 users) mentioned ADHD, 24,105 tweets (7224 users) mentioned ASD, 2957 tweets (1515 users) mentioned delayed speech, and 5226 tweets (3078 users) mentioned asthma. We used 400 matching tweets—100 tweets per outcome—to develop annotation guidelines to help three annotators distinguish tweets that reported having a child with ADHD, ASD, delayed speech, or asthma, from those that did not. Then, an additional 9334 tweets—one random tweet per user—were independently annotated, with 8334 tweets being dual annotated and 1000 tweets being annotated by all three annotators. Among the 9734 total annotated tweets, 1671 mentioned ADHD, 5220 mentioned ASD, 995 mentioned delayed speech, and 2170 mentioned asthma, with some tweets mentioning multiple disorders.

### Automatic classification

We used the 9734 annotated tweets in supervised machine learning experiments to train and evaluate binary classification. For the classifiers, we used the LibSVM [19] implementation of support vector machine (SVM) in Weka and three deep neural network classifiers based on bidirectional encoder representations from transformers (BERT): the BERT-Based-Uncased [20], RoBERTa-Large [21], and BERTweet-Large [22] pretrained models in the *Flair* Python library. We split the tweets into 80% (7787 tweets) and 20% (1947) random sets as training data and held-out test data, respectively. For the SVM classifier, we preprocessed the tweets by normalizing URLs, usernames, names, digits, and references to ADHD, ASD, delayed speech, asthma, and a child, removing non-alphanumeric characters and extra spaces, and lowercasing and stemming [23] the text. We used Weka’s NGram Tokenizer to extract n-grams (n = 1-3) as features in a bag-of-words representation. We used the radial basis function kernel and set the *cost* at *c* = 50 and the class weights at *w* = 1 for the “negative” class and *w* = 2 for the “positive” class. For the BERT-based classifiers, we preprocessed the tweets by normalizing URLs and usernames, and lowercasing the text. We used Adam optimization, a batch size of 8, 5 epochs, and a learning rate of 0.00001. We fine-tuned all layers of the transformer models with our annotated tweets.

## RESULTS

Based on 1000 tweets that were independently annotated by all three annotators, inter-annotator agreement (Fleiss’ kappa) was 0.88. After resolving the disagreements among all 9734 annotated tweets, we determined that 3019 (31%) tweets reported having a child with ADHD, ASD, delayed speech, or asthma, and 6715 (69%) did not. Specifically, 382 (23%) of the 1671 tweets that mentioned ADHD reported having a child with ADHD; 1265 (24%) of the 5220 tweets that mentioned ASD reported having a child with ASD; 584 (59%) of the 995 tweets that mentioned delayed speech reported having a child with delayed speech; and 926 (43%) of the 2170 tweets that mentioned asthma reported having a child with asthma.

Table 1 presents the precision, recall, and F_1_-score of SVM and deep neural network classifiers for the class of tweets that reported having a child with ADHD, ASD, delayed speech, or asthma, evaluated on a held-out test set of 1947 (20%) of the 9734 annotated tweets. The classifier based on a RoBERTa-Large pretrained model achieved the highest overall F_1_-score: 0.93 (precision = 0.92 and recall = 0.94). Table 1 also presents the precision, recall, and F_1_-score of the RoBERTa-Large classifier for tweets that mention specific outcomes. We deployed the RoBERTa-Large classifier on the additional 26,360 unlabeled tweets that matched our query. Between the 9734 manually annotated tweets and the 26,360 automatically classified tweets, we identified 3806 total users who reported having a child with ADHD (678 users), ASD (1744 users), delayed speech (902 users), or asthma (1255 users).

**Table 1.**
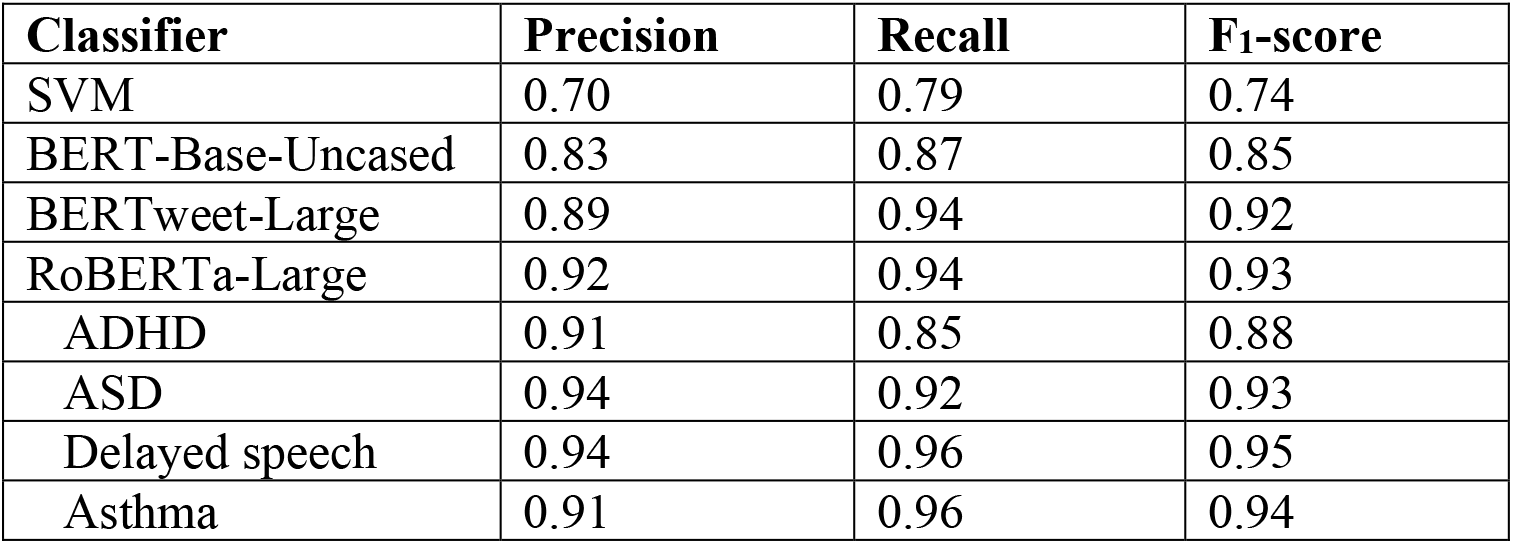
Precision, recall, and F_1_-score of classifiers for detecting tweets that report having a child with attention-deficit/hyperactivity disorder (ADHD), autism spectrum disorders (ASD), delayed speech, or asthma, including outcome-specific precision, recall, and F_1_-score for the RoBERTa-Large classifier.

Table 2 presents examples of false positives and false negatives of the RoBERTa-Large classifier in the test set. Among the 48 false positives, 28 (58%) do refer to the user’s child; however, among these 28 false positives, 11 (39%) indicate that someone other than the user’s child—for example, the user, a family member, or children in general— has ADHD, ASD, delayed speech, or asthma (Tweet 1), 9 (32%) indicate that a disorder is merely suspected or exhibited (Tweet 2), and 4 (14%) indicate that the user’s child has a disorder that is not relevant to this study (Tweet 3). Among the other 20 (42%) of the 48 false positives, 10 (50%) are reported speech, such as quotations (Tweet 4), 5 (25%) indicate that the user is not a biological parent—for example, a guardian, educator, carer, or relative—of a child with ADHD, ASD, delayed speech, or asthma (Tweet 5), and 4 (20%) refer to someone with a disorder, but are ambiguous about whether that person is the user’s child (Tweet 6). Among the 42 false negatives, 22 (52%) do not explicitly mention the user’s child (Tweet 7)—for example, referring to the child with a pronoun or name—and 14 (33%) do not explicitly indicate that the child has ADHD, ASD, delayed speech, or asthma (Tweet 8). Among the 31 (74%) false negatives that do not explicitly (1) mention the user’s child or (2) indicate that the child has one of the disorders, 6 (19%) mention another person who also has the disorder (Tweet 9). Among the other 11 (26%) of the 42 false negatives, an additional 6 (55%) mention another person who also has the disorder (Tweet 10).

**Table 2.** Sample false positives and false negatives of a RoBERTa-Large classifier for detecting tweets that report having a child with attention-deficit/hyperactivity disorder (ADHD), autism spectrum disorders (ASD), delayed speech, or asthma, with the text that matched our data collection query in bold.

|  | **Tweet** | **Actual** | **Predicted** |
| --- | --- | --- | --- |
| 1 | So Maxine Waters can be maskless on a plane but **I** can’t fly with **my 2 year old** cause she won’t wear a mask? **Kids** with **autism** are being banned from flying because they won’t wear a mask? | - | + |
| 2 | they treat **my baby** with **asthma** meds all the time but didn’t diagnose her with it im pretty sure she has it tho | - | + |
| 3 | [name] has Williams Syndrome, not **autism** but the other day someone said “**my** heart breaks for you”.. and **I** had to explain but things might not be the easiest or ideal but he s the happiest **baby** and is loved by everyone that knows him :) | - | + |
| 4 | Any tips for this mum: " **My daughter** is 10. **My** parents would like to gift her either a phone or a smart watch which is easy to use and won't be easily damaged by a very active **ADHD kid**... I need help choosi... [URL] | - | + |
| 5 | [name] is **autistic** & I couldn't for a second be annoyed of his tantrums or his unique ways . I love that **kid** like **my** own, & I just understand him always. | - | + |
| 6 | If **we** were all healthy and vaccinated I'd be all for sending them but **we** have two diabetics, an **asthmatic**, and a **toddler**. We're stuck. | - | + |
| 7 | Flying tomorrow...during a pandemic with a **nonverbal 3 year old**. We could use some prayers, please. 😞 😞 | + | - |
| 8 | I wouldn't change **my child** for anything in the world. I'm just curious to know where **autism** came from because **me** and his dad don't have any family members that are **autistic**. It's just a question out of curiosity | + | - |
| 9 | Know that instead of your world falling apart, you just got more information about your **kid** and now you can be an even better **parent**! Follow other **autistic** people on here and ask a million questions. That's what I do. | + | - |
| 10 | I've kept **my daughter** off nursery since last week as herself and I are both **asthmatic** and I just don't want to risk **my child's** health. | + | - |

## DISCUSSION

Our ability to identify Twitter data during pregnancy for users who reported having a child with ADHD, ASD, delayed speech, or asthma demonstrates that Twitter could be a complementary source of data for assessing associations between pregnancy exposures and childhood health outcomes. In our recent work, we were able to identify prenatal, perinatal, and postnatal outcomes—miscarriage, preterm birth, low birthweight, birth defects, and NICU admission—for Twitter users who posted tweets reporting that they took medication [11] or received COVID-19 vaccination [12] during pregnancy. The results of the present study suggest that the use of Twitter data for monitoring drug safety and COVID-19 vaccine safety in pregnancy could be extended to outcomes that are typically beyond the follow-up period of pregnancy registries, such as the *V-safe COVID-19 Vaccine Pregnancy Registry* [24]. As we continue collecting users’ tweets and adding users to our cohort in real-time [17], our ability to automatically detect these outcomes with an overall F_1_-score of 0.93 and outcome-specific F_1_-scores between 0.88 and 0.95 demonstrates the feasibility of using Twitter data for observational studies on a large scale.

## CONCLUSION

In this paper, we presented a manually annotated data set demonstrating that there are users who were active on Twitter during pregnancy and also posted tweets that reported having a child with ADHD, ASD, delayed speech, or asthma. The performance for automatically identifying these outcomes (F_1_-score = 0.93) enables large-scale observational studies, advancing the use of Twitter data for epidemiology of pregnancy outcomes for which the causes are largely unknown and sources of data are limited.

## Data Availability

All data produced in the present study are available upon reasonable request to the authors.

## FUNDING

This work was supported by the National Library of Medicine (R01LM011176).

## AUTHOR CONTRIBUTIONS

AZK and JAGG: data collection, annotation, machine learning experiments, error analysis, and writing the manuscript. LDL: providing guidance on pregnancy outcomes and editing the manuscript. GGH: designing and guiding the study, and editing the manuscript.

## ACKNOWLEDGMENTS

The authors thank Ivan Flores for contributing to software applications, and Karen O’Connor for contributing to annotating the Twitter data.

## CONFLICT OF INTEREST STATEMENT

None declared.

## Notes

### Competing Interest Statement

The authors have declared no competing interest.

### Author Declarations

This study used publicly available Twitter data. The Institutional Review Board of the University of Pennsylvania reviewed this study and deemed it exempt human subjects research under 45 CFR 46.101(b)(4) for publicly available data sources.

